# The Effectiveness of Using a Clinical Support Tool in Managing Adolescents with Non-Traumatic Knee Pain (MAP-Knee): protocol for a cluster-randomised superiority trial with delayed intervention and embedded realist evaluation

**DOI:** 10.1101/2024.04.29.24306576

**Authors:** Henrik Riel, Simon Kristoffer Johansen, Erika Maria Andersen, Malene Kjær Bruun, Niels Henrik Bruun, Chris Djurtoft, Simon Doessing, Tina Heyckendorff-Diebold, Per Hölmich, Martin Bach Jensen, Søren Kaalund, Niels-Christian Kaldau, Tommy Frisgaard Oehlenschlaeger, Charlotte Overgaard, Ole Rahbek, Dorthe Brøndum Rasmussen, Susanne Olesen Schaarup, Thomas Sørensen, Louise Lund Holm Thomsen, Michael Skovdal Rathleff

## Abstract

**Background:** Knee pain affects one in three adolescents, which makes it one of the most common pain sites. Guideline recommendations about the clinical selection of patients likely to benefit from interventions are unclear, which leads to treatment heterogeneity and the potential of wasted resources among adolescents with a good prognosis. In contrast, adolescents with a poorer prognosis may not receive sufficient care. A newly developed clinical decision-support tool (The MAP-Knee Tool) intends to support clinicians in engaging with patients and adjusting the clinicians’ evidence-based practices to accommodate patient preferences and treatment needs via a shared decision-making process. The aims of this trial are 1) to investigate the effectiveness of using a clinical decision-support tool (The MAP-Knee Tool) compared with usual care in adolescents with non-traumatic knee pain in reducing pain measured by KOOS-Child Pain after 12 weeks and 2) to investigate how the intervention worked, for whom, why and under which circumstances applying realist evaluation methodology.

**Methods:** This trial is a cluster-randomised superiority trial with a delayed intervention and a realist evaluation. Six hospital departments start with a usual care period of 4 months before randomly crossing over to using the intervention (The MAP-Knee Tool) after 4, 6, or 8 months, respectively. We will recruit 290 adolescents suffering from non-traumatic knee pain diagnoses who are followed for one year, with the change in KOOS-Child Pain after 12 weeks considered the primary endpoint. Secondary outcomes include 1) Global Rating of Change, 2) EQ-5D-Youth, 3) Anterior Knee Pain Youth, 4) the International Physical Activity Questionnaire short version, and 5) sports participation. The realist evaluation will utilise a prospective, qualitative approach for collecting data needed to develop and test a program theory and identify context-mechanism-outcome configurations essential for understanding how outcomes are achieved within specific contexts.

**Discussion:** This trial focuses on how the initial clinical encounter can be improved to meet the support and management needs of adolescents with chronic knee pain seeking treatment for knee pain in secondary care and investigates how the intervention worked, for whom, why and under which circumstances.

**Trial registration:** Clinicaltrials.gov (NCT05791513). Prospectively registered on March 30th, 2023.

## Background

From the age of 5–9 years to the age of 10–14, there is an almost fourfold increase in the years lived with disability as a result of musculoskeletal conditions (1). In Danish primary care, there is an eightfold increase in contacts in our healthcare sector because of knee symptoms between 5–9 and 10–19 years of age and approximately half of these adolescents are referred to the secondary care sector (2,3). This makes knee pain one of the most common pain sites experienced by youths and adolescents and can affect their school attendance, participation in hobbies and social activities, and mental health (4–6). The most common non-traumatic knee pain diagnoses are Patellofemoral Pain and Osgood-Schlatter Disease; however, patellar tendinopathy, Sinding-Larsen Johansson, and iliotibial pain syndrome are frequent differential diagnoses.(7–9)

Knee pain affects one in three adolescents, and around 60 % will seek medical care due to knee pain (10). Data from prospective cohorts show that 40-50% will still experience knee pain and symptoms after 2-5 years (11–13). A cohort study from our group documented that 70% of adolescents with Patellofemoral Pain reduced or stopped participating in sports during a two-year period, and they had significantly worse EQ-5D Index and VAS scores than those without knee pain (12). This underlines the need for effective treatments to prevent long-term disability and physical inactivity.

There are several management strategies for adolescents suffering from knee pain. These range from minimal support, including patient education and leaflets regarding self-management, to specialised supervised exercise therapy (14–17). Because of insufficient evidence, guideline recommendations about the clinical selection of patients likely to benefit from referral to additional interventions are unclear, which leads to treatment heterogeneity (3,11). Therefore, resources may be wasted among adolescents with a good prognosis, while adolescents with a poorer prognosis may not receive sufficient care. Patient decision aids can support clinicians in engaging with patients and adjusting their evidence-based practices to accommodate patient preferences and treatment needs via a shared decision-making process. Such aids may be especially relevant in conditions with several treatment options available and thus may improve care for adolescents suffering from non-traumatic knee pain.(18–20)

We recently developed a clinical decision-support tool intending to support shared decision-making between adolescents, medical doctors, and parents. The development included systematic literature searches, an Argumentative Delphi process, and end-user involvement (21), ultimately leading to the MAP-Knee Tool (22), which clinicians may use during consultation with adolescents with non-traumatic knee pain. The tool is intended to support the entire consultation, from diagnosing the condition and explaining the condition to providing information regarding possible management options. The feasibility of using the tool in a clinical setting has been established (Bruun et al. *in preparation*), but whether the MAP-Knee Tool is associated with improved long-term outcomes among adolescents with non-traumatic knee pain compared to usual care is unknown.

The overall aims of this trial are 1) to investigate the effectiveness of using our novel clinical decision-support tool compared with usual care in adolescents with non-traumatic knee pain in reducing pain measured by KOOS-Child Pain after 12 weeks and 2) to investigate how the intervention worked, for whom, why and under which circumstances using a qualitative realist evaluation (23). We hypothesise that adolescents with non-traumatic knee pain who have been consulting a clinician who uses the MAP-Knee Tool will be superior in self-reported recovery and physical function after 12 weeks compared to adolescents who have been consulting a clinician not using the MAP-Knee Tool.

## Methods

### Design and setting

The trial is designed as a cluster-randomised superiority trial with a delayed intervention and a realist evaluation component (23). All hospitals start with a control period of 4 months before randomly crossing over to using the intervention after 4, 6, or 8 months. The trial is conducted across six hospital departments in Denmark (Aalborg University Hospital, Amager-Hvidovre Hospital, Bispebjerg Hospital, North Denmark Regional Hospital Frederikshavn, Næstved Hospital, and Vejle Hospital). Future reporting of the trial will follow CONSORT guidelines for reporting non-pharmacologic treatments (24). Reporting of this protocol follows the SPIRIT statement (25). The trial was planned in accordance with the PREPARE Trial guide (26). Before the inclusion of the first participant, the trial was registered on clinicaltrials.gov (NCT05791513), and the trial was approved by the Ethics Committee of the North Denmark Region (N-20220043).

The design of the realist evaluation was anchored in the realist evaluation circle to facilitate theory generation, which included formulating an initial theory of how the intervention brings about change, generating hypotheses about essential mechanisms and processes, collection and analysis of empirical data and program theory testing (23,27). The realist evaluation will involve several data collections conducted before, during and after the stepped wedge trial to support theory gleaning, refinement and consolidation (28). Data will be collected via document analysis, realist interviews with researchers who developed the MAP-Knee Tool, clinicians who will be implementers of the MAP-Knee Tool and adolescents with non-traumatic knee pain who received treatments with the MAP-Knee Tool. The data will be analysed prospectively using the linked coding approach by Jackson and Kolla (29) to identify context-mechanism-outcome (CMO) strings within the coded texts and Gilmore et al.’s (30) memo-coding technique to extract CMO configurations from our data and inform the refinement and consolidation of the intervention program theory (23,31). The program theory will synthesise insights from the document and realist interviews with insights from the quantitative data collected at the 12-week follow-up (27,32) and evaluated by clinicians using Mukumbang et al.’s (33) approach for including program theory segments during realist interviews to obtain feedback and consolidate program theories and context-mechanism-outcome configurations.

Adolescents with non-traumatic knee pain will be recruited when they arrive at one of the six hospitals due to a referral from their general practitioner. In contrast, clinicians and adolescents with non-traumatic knee pain for the realist evaluation will be included prospectively based on information power before, during and after the initiation of the MAP-Knee trial (34). Thus, recruitment is based on the natural flow of patients with non-traumatic knee pain seen at the hospital and the availability of clinicians who have used the MAP-Knee Tool in their treatment of adolescents with non-traumatic knee pain. Adolescents will only attend the baseline examination at the hospital, whereas a link to questionnaires is emailed to them for the 12-week and 52-week follow-ups using REDCap (Vanderbilt University, Nashville, TN, USA). They may have additional appointments at the hospital during the trial, depending on the treatment plan, but they will not answer questionnaires during these clinical encounters.

### Roles and responsibilities

The project manager is an experienced physiotherapist who coordinates the trial. When potential participants arrive at the hospital, they are informed about the trial by either a project nurse or project secretary. This person collects written informed consent from participants and their parents/legal guardians if the participant is under the age of 18 years and collects data using questionnaires. After this, the participant is seen by a clinician (orthopaedic surgeon, physiotherapist, rheumatologist, general practitioner). The type of clinician varies from one hospital to another due to differences in how each hospital is organised. The realist analysis will be led by a researcher with a background in Information Science and extensive experience in planning and conducting qualitative studies with youths with chronic pain and healthcare providers, supported by experts in conducting realist evaluations in healthcare settings. He is supervised by experts in conducting realist evaluations (CO and LLH).

### Eligibility criteria

The inclusion criteria for the adolescents are: 1) having been referred to a hospital due to non-traumatic knee pain and 2) being aged between 10 and 19 years. Exclusion criteria: 1) knee pain with a traumatic origin or 2) lack of ability to cooperate. Any orthopaedic or similar hospital department in Denmark to which adolescents suffering from non-traumatic knee pain may be referred was eligible for participation. Any medical doctor or physiotherapist employed at either of the study sites who regularly sees adolescents with non-traumatic knee pain is eligible to participate in the trial. The inclusion criteria for the researchers participating in the realist evaluation were: 1) having participated in the conceptualisation, development or testing of the MAP-Knee Tool, or 2) any of the relevant sub-studies, or 3) planning of the MAP-Knee Trial. Finally, the inclusion criteria for clinicians included in the realist evaluation were 1) being a healthcare provider (e.g., physiotherapist, orthopaedic surgeon, rheumatologist) who is 2) employed at one of the study sites, and 3) either had or was likely to use the MAP-Knee Tool when treating adolescents with non-specific knee pain.

### Interventions

Before crossover occurs at the hospital, the adolescents will be diagnosed and treated as usual at the treating clinician’s discretion. Based on our previous research on the usual care pathway, this will be heterogeneous and include advice to wait and see, imaging (most often MRI and x-ray), or a rehabilitation plan for treatment within the municipality setting (3,11).

After crossing over to using the MAP-Knee Tool, the treating clinician will use the MAP-Knee Tool together with the adolescent. The tool was designed to support the entire consultation, from diagnosing the condition (Patellofemoral Pain, Osgood-Schlatter, Sinding-Larsen-Johansson, growth pain, patellar tendinopathy, or iliotibial band syndrome) to deciding on future management. The MAP-Knee Tool includes four separate components: 1) a tool for diagnosing the most common types of non-traumatic knee pain (SMILE) (35), 2) credible explanations of the aetiology and pathogenesis specific to the diagnosis based on multiple methods with an iterative design, 3) a presentation of prognostic factors based on an individual participant data meta-analysis(36), and 4) an option grid that presents the users of the tool with pros and cons of commonly used management options, derived from a systematic literature search of systematic and narrative reviews within non-traumatic adolescent knee pain. An overarching focus of all components was to support shared decision-making and base decisions on all three pillars of evidence-based medicine: patient values, clinical expertise, and relevant research.(37) Therefore, the tool should not provide clinicians using the MAP-Knee Tool with definitive answers simply based on the available evidence. After the initial prototype of the tool, including all four components, had been made, we performed end-user testing using think-aloud sessions with adolescents suffering from non-traumatic knee pain, adolescents with no history of knee pain, and medical doctors.(22)

Before the start of crossover, there will be a 2-week transition period for training the clinicians in using the support tool in their clinical practice. The clinicians will participate in a 1-hour training session with the project manager, where they will be educated on using the MAP-Knee Tool. After this session, they will be asked to use the tool for one week in clinical practice, and a follow-up session will be held where they can share their experiences with using the tool and ask the project manager questions. Then, the clinicians can use the MAP-Knee Tool for another week before the patients seen with the tool are included in the trial. The patients seen during the 2-week transition period will not be included in the trial. To improve long-term adherence to using the tool, there will be a booster session similar to the follow-up after the first week in the transition period after four weeks.

The research activities related to the realist evaluation will run in parallel with the activities of the MAP-Knee Trial while moving from the initial theory-gleaning phase and formulating preliminary program theories to phase 2, which entailed the data collection during the crossover of hospital department 1 and 2 (23). The realist evaluation will utilise a prospective, qualitative approach for collecting the qualitative data needed to develop and test a program theory and identify context-mechanism-outcome configurations (23), delineating the micro, meso, and macro-level barriers and facilitators for archiving positive health outcomes from interventions with the MAP-Knee Tool (38).

### Documentation analysis

All available documents (protocols, logic models, pre-preprints, peer-reviewed studies) will be collected, categorised, and analysed through a realist thematic analysis approach by Wiltshire and Ronkainen (39) using NVivo 14 coding software to identify thematic patterns across texts and using this insight to glean the underlying causal mechanisms which researchers envisioned would facilitate behavioural change and tool acceptance during the conceptualisation and design of the MAP-Knee Tool.

### Realist interviews

We will conduct realist interviews with relevant stakeholders using the principles for planning and conducting realist interviews by Manzanos (28) and Mukumbang et al. (31) to maintain theoretical awareness during initial theory gleaning and subsequent theory testing interviews. Theory-gleaning interviews with the researchers behind the MAP-Knee Tool will be conducted before the first departments cross over, using an interview guide with open and probing questions informed by the document analysis. Additionally, one realist interview will be conducted with a clinician (physiotherapist, orthopaedic surgeon, rheumatologist) during the time of the crossover to identify the clinicians’ expectations, thoughts, needs, and contextual challenges related to using the MAP-Knee tool when treating adolescents with non-specific knee pain. The theory-gleaning interviews will inform the initial program theory (IPT). The initial theory will be tested and expanded during the intervention with the MAP-Knee tool. The testing will be conducted through (n=3) realist interviews with clinicians (physiotherapists, orthopaedic surgeons, rheumatologists) per department, and (n=6-8) adolescents with knee pain, who received treatment with the MAP-Knee Tool across the timespan of the intervention. By exploring how clinicians and adolescents with non-traumatic knee pain experienced the MAP-Knee Tool facilitated the treatment of non-traumatic knee pain, we will test, revise and expand the initial program theory and identify the CMO configurations present within the use situation.

To ensure the integrity of the contents of the program theory and the identified CMO configurations, the program theory will be tested and validated through realist interviews with selected clinicians from each department using the program theory to test and consolidate findings (33). The realist interviews will test, revise, and finalise the program theory to ensure no significant points are missed during the data collection and analysis. This will ensure the program theory’s relevance, scalability, and transferability and validate the identified CMO configurations.

### Randomisation

Hospitals are randomly allocated to start the interventional period after 4, 6, or 8 months, respectively, using a random list generator (www.random.org). The project manager generated the list and will inform the hospitals when they should cross over as late as possible but no later than one month before the crossover.

### Variables

#### Descriptives

During baseline, we will collect the following data: age, height, weight, location (unilateral or bilateral) and duration of the knee pain, pain frequency, pain intensity during the week prior to baseline measured on a 0 to 10 Numerical Rating Scale (0=no pain, 10= worst pain imaginable), and pain medication use. At each follow-up, adolescents will be asked which treatment they have received and which clinicians they may have consulted.

#### Primary outcome

The primary outcome will be change in KOOS-Child pain subscale (40) from baseline to the 12-week follow-up. KOOS-Child consists of 39 items divided into five subscales (Pain, Symptoms, Activities of Daily Living, Sport/Recreation, and Quality of Life) ranging from 0 to 100, with 100 being optimum, and we use a Danish translation of the questionnaire. This questionnaire has been found to have good psychometric properties and is recommended for children and adolescents suffering from knee disorders.(40) The questionnaire will be completed at baseline and the 12-week and 52-week follow-ups.

#### Secondary outcomes

Secondary outcomes include: 1) Global Rating of Change (GROC), 2) EQ-5D-Youth, 3) Anterior Knee Pain Youth (AKP-Youth), 4) the International Physical Activity Questionnaire short version (IPAQ), and 5) sports participation.

We will use the GROC to measure the participants’ self-reported recovery on a 7-point Likert scale ranging from “much improved” to “much worse”. Participants are categorised as improved if they rate themselves as “much improved” or “improved” (category 6-7) and categorised as not improved if they rate themselves from “slightly improved” to “much worse” (category 1-5). Health-related quality of life will be estimated by the EQ-5D-Youth questionnaire, an adapted version of the EQ-5D used in an adult population. Yet, it still consists of the same five subscales, which are mobility, self-care, usual activities, pain and discomfort, and anxiety and depression.(41) We will use our newly developed questionnaire (AKP-Youth) as a secondary measure of condition severity. The AKP-Youth contains 23 items that are divided into four overarching domains of impact: symptoms, limitations in physical activity, limitations in social activities, and emotional impact of pain. Changes in physical activity level will be measured by the IPAQ. A Danish translation of the original questionnaire will be used. The IPAQ is the most used questionnaire for measuring physical activity and consists of 9 items that provide information on the time spent performing vigorous and moderate activities, the time spent walking, and the time spent sedentary during the past week. The IPAQ estimates the total weekly physical activity measured in MET minutes per week and the total minutes spent sitting.(42,43) Change in sports participation will be explored using a questionnaire in which participants are asked whether they have participated in leisure sports activities, type of sports activities and weekly frequency. All questionnaires will be completed at baseline and at the 12-week and 52-week follow-ups. The GROC is only being answered during the 12-week and 52-week follow-ups.

### Adverse events

The participants will be able to report any adverse events to the primary investigator when they occur by phone, SMS, or e-mail. Adverse events will be graded 1 to 5 according to the Common Terminology Criteria for Adverse Events v4.03 (44). A medical doctor will assess and grade the adverse event. Participants will not be withdrawn from the trial due to an adverse event. If a participant experiences an adverse event and requests withdrawal from the study, data until the adverse event occurred will be included in the analyses. The project manager will report any incidents to the sponsor as quickly as possible and no later than 15 days after the participant reports the event. Sponsor will report any severe adverse events (grade 3-5) to the Ethics Committee of North Denmark Region no later than seven days after being informed.

### Sample size

#### Trial

The trial is powered to detect a between-group difference of at least 10 points on the KOOS-Child questionnaire pain subscale scale (0 to 100) (40). Based on a standard deviation of 22 points, type I error α =0.05 and power = 0.90, a conservative estimate of four steps and an intraclass correlation coefficient = 0.01, we will need at least 240 participants (20 per step per cluster). We will increase the sample size to 290 participants across the six clusters to account for a larger outcome variance.

#### Realist Evaluation

Participants for the realist evaluation will be selected progressively based on an ongoing assessment of participants’ knowledge power (34) and how their experiences and knowledge inform the theory-gleaning, testing and consolidation phases of the realist evaluation (45). We anticipate that 15-20 clinician interviews and 4-8 interviews with adolescents with non-traumatic knee pain treated with the MAP-Knee Tool will be sufficient to inform the development, testing and consolidation of a program theory and ensure data completeness throughout the project.

### Statistical analyses

All statistical analyses will be performed according to a pre-established analysis plan by a statistician. This plan is written with the statistician and will be published on the Aalborg University website before the inclusion of the last participant. Stata ver. 18 will be used as statistical software. We will use Q-Q plots and histograms to assess data distribution. The primary intention-to-treat analysis will investigate the between-group difference in KOOS-Child pain at the 12-week follow-up using a linear mixed effects model with the participant as random effect. The baseline value, time (12 and 52 weeks), group allocation (using or not using MAP-Knee Tool) and term for the interaction between time and group will be treated as fixed-effect variables. We will apply the same model for the other continuous outcomes. The relative risk (RR) will be calculated for the dichotomised GROC to determine the probability of being improved after 12 and 52 weeks. The number needed to treat will be calculated as 1/risk difference.

### Qualitative data analysis

The data collected from the realist interviews with researchers, clinicians, stakeholders, and adolescents with non-traumatic knee pain will be collected, stored, and analysed prospectively using Gilmore et al.’s (30) memo-coding technique for extracting CMO configurations from qualitative data. The memo-coding technique was selected because it focuses on extracting CMO configurations and incorporating them into a program theory through a five-step process, which included data preparation, CMO configuration extraction, program theory development, refinement, and synthesis across multiple data collections. The data analysis will be conducted using NVivo 14 to sort and organise the data. The linked coding approach by Jackson and Kolla (29) will be used to identify CMO strings within the raw data before using the memo-coding technique to derive CMO configurations from the data. By adopting the memo-coding technique for organising and synthesising our data, we obtain a foundation for bridging the gap between identifying ‘empirical themes’, exploring ‘co-occurring phenomena’ within the data and extracting the ‘structures and mechanisms’ which generate the actual phenomena present within the use situation and informing our program theory (30,33).

The analysis will be conducted in steps by two researchers (SKJ; EMA) and involve the uptake and multiple datatypes (document, researcher, clinician, clinical staff, adolescents) to inform the ongoing development of the program theory (23,27,32). The data preparation and transcription will be conducted using Whisper 1.1 (Open AI, San Francisco, CA, USA) AI transcription software and ExpressScribe software v. 7.01 (NCH Software, Canberra, Australia) to check and resolve transcription errors within the data before analysis. The text files from the document-, researcher-, clinician and validation interviews will be uploaded and analysed sequentially using NVivo 14 analysis using the following steps.

The initial transcription of interview data will be undertaken using the Whisper 1.1 tool. The AI-transcribed files will be checked manually by a researcher using ExpressScribe 7.05 transcription software to ensure transcript integrity. Before the analysis, the preliminary CMOs from the initial program theories (IPT) gleaned during the document analysis were entered as nodes within NVivo 14. During the data coding, potential CMOs will be identified via the linked coding approach and added to an existing node (IPT) or a new node linked to an existing node (30). All nodes with encoded data will be reviewed and subjected to interpretation using linked memos to summarise the contents, extract CMO configurations and identify their internal relationships. From this, the extracted CMO configurations will be organised hierarchically, using the child nodes to evaluate, support, refute, revise, and expand the IPTs through a memo writeup process. The revised IPTs and CMO configurations from each analysis will be integrated within a logic model (33) to inform the formulation of a program theory as the analysis progresses, to identify the change mechanisms acting as barriers and facilitators for supporting patient education, stratification and shared decision-making when using the MAP-Knee Tool in complex treatment settings.

### Validation interviews

The contents of the program theory and identified CMO configurations will be tested and subjected to evaluation through realist interviews with selected clinicians from each department who had used the MAP-Knee Tool when treating adolescents with non-traumatic knee pain. The validation interviews will combine principles from realist interviewing using selected parts of the program theory to inform discussions (31) to solicit participants’ feedback and suggestions for optimising the program theory and CMO configurations to reflect their experiences by using the MAP-Knee Tool, and the individual and contextual barriers and facilitators for supporting patient education, stratification, and shared decision-making.

### Data monitoring and quality assurance

All data will be stored electronically and handled according to the General Data Protection Regulation. Data safety may be overseen unannounced by the Danish Data Protection Agency. Participant data will be stored in REDCap, whereas data processor agreements, collaboration agreements between the project group and hospitals, and protocols will be stored on a secure server at Aalborg University. Data collection instruments have been developed in REDCap to prevent data entry errors so that required data must be included or an error will be displayed. Validation of each field has been chosen (e.g., if the data format does not appear to be a date in the field ‘Date’, an error is displayed). Data are checked once per week by the project manager to ensure no missing data, and participants and/or their parents are contacted if they do not respond to the questionnaires sent to them for the 12-week and 52-week follow-ups. All data will be kept for ten years after the trial is completed per the European Code of Conduct for Research Integrity.

## Discussion

### Moving from “what we prescribe” to “how it is delivered”

Previous trials to support adolescents with chronic knee pain have focused on what clinicians should deliver (e.g., exercise, orthoses, taping or load management (14,16,17,46,47)). This trial focuses on how the initial clinical encounter and clinicians’ decision-making can be improved to meet the support and management needs of adolescents with chronic knee pain seeking treatment for knee pain in secondary care. Previous studies have highlighted how adolescents’ formation of self-management strategies does not occur within a vacuum (48) and how the lack of validation from HCPs, peers and parents, uncertainty about the severity of the condition, excessive use of diagnostic imaging and ‘watchful waiting’ without explanations may negatively influence adolescents’ formation of strategies for mastering their knee pain (49–52). Contrarily, patients highlight how validation from clinicians, a name, and an explanation for why their knee pain emerged enabled adolescents to adjust their pain beliefs and commence the work related to accepting and exploring how to self-manage their knee pain in everyday situations (52). Studies from other domains have described how informed parents can help facilitate adolescents’ transition into self-management by providing comfort, management instructions, problem-solving assistance and engaging with external actors (teachers, trainers, etc.) (53,54), adolescents with knee pain and parents described how they needed different types of information from HCPs to undertake this task (51). Whereas shared decision-making has been highlighted as a viable method for merging evidence-based treatments and patients’ desired outcomes when prescribing treatments in clinical settings (55), the question of how patient education is delivered to enable adolescents and parents to engage in an informed negotiation of roles and management tasks is unexplored (56). Thus, we anticipate that the inclusion of the MAP-Knee Tool can help shift the clinicians’ focus from simply prescribing treatments to how treatments and patient education can be delivered to make it ‘actionable’ for adolescents with knee pain by visualising the clinicians’ decision-making process, similar to what Star (57) describes within their boundary object—ensuring that clinicians understand how the MAP-Knee Tool is not a treatment in itself but rather a tool for merging their existing clinical expertise with patient-centred care principles via a systematised approach.

### A tool that can be refined when new evidence emerges

The MAP-Knee Tool should not be viewed as a static tool but as a tool that can be updated when new evidence emerges. This will ensure that clinicians have information regarding best practice readily available to them. Therefore, it may be preferable to convert the tool into an online version in the future. The current physical form of the tool is not seen as crucial as it is merely a delivery vehicle of the content, which is the fundamental aspect of the MAP-Knee Tool. Yet, using a leaflet can extend the clinical encounter, and the use of written materials has been found to aid patients with chronic pain conditions in reducing pain catastrophising.(58)

### Limitations of the trial design

An important potential limitation of the trial is the nature of the design in which recruitment relies on the natural flow of patients referred to the hospitals. This does not allow adjusting the recruitment strategy if the expected recruitment rate is unmet. Based on our past experiences when recruiting adolescents with non-traumatic knee pain, the recruitment rate required to achieve the sample size was feasible. However, due to a lower-than-expected recruitment rate during the first two months of recruitment, we had to postpone the first crossover from after two months to after four months of using usual care. To balance the periods of all hospitals using usual care with all hospitals using The MAP-Knee Tool, we also extended the period after the last crossover from two to four months. Due to the learning curve when starting to use the tool, it may be challenging for clinicians to become confident in using it if the recruitment rate is not somewhat consistent. This could potentially limit the implementation. Another challenge that could hamper recruitment is the fact that the ethics committee did not approve only obtaining informed consent from a single parent/legal guardian. Therefore, we must obtain retrospective informed consent from the other parent in cases where only one parent attends the clinical examination at the hospital. If the other parent cannot be contacted or does not want to sign the informed consent form, the participant will need to be excluded.

### Moving from clinical trials to understanding “how and why things work”

While multiple studies have explored implementing decision-making tools to support the cultivation of patient-centred and collaborative care approaches in primary and secondary care settings (59–62), these studies commonly focus on measuring the effects of interventions. This is problematic, as studies from implementation science highlight how care settings are complex, dynamic, and adaptive systems (63) and how introducing a tool for, e.g. optimising HCP workflows, may ripple into other domains and influence the tasks, roles, division of labour and community and cultural aspects within cares setting (64,65). Furthermore, introducing tools may also have unforeseen impacts on workflows, collaborations, and communications within the organisations, resulting in implementers having to use extra resources to find workarounds when using the tool (66). Thus, the historical and traditional focus on exploring the effect of interventions has resulted in a dearth of knowledge on how future treatment and supporting tools should be modelled to support their meaningful integration into complex treatment settings. This is also true in Danish secondary care. By adopting a realist perspective on the testing and evaluating the MAP-Knee Tool, an optic is obtained that enables us to transcend the scope of the clinical trial and explore how, why, and which circumstances result in the outcomes of the MAP-Knee Trial (23). Furthermore, by adapting the view of interventions as programs as described by Pawson and Tilley (23), the qualitative and quantitative data collected during the intervention with the MAP-Knee Tool can be synthesised to identify the causal powers influencing the implementation of the tool on a micro, meso, and macro level. By identifying the different context mechanisms activated by the MAP-Knee Tool intervention, visualised within a program theory, the study has the potential to extract novel, general insights on why some interventions fail and for whom, while others succeed, of high relevance for future studies in secondary care.

## Data Availability

All data produced in the present study are available upon reasonable request to the authors

